# Outcomes of NAFLD and MAFLD: Results from a community-based, prospective cohort study

**DOI:** 10.1101/2020.09.23.20200535

**Authors:** Madunil Anuk Niriella, Dileepa Senajith Ediriweera, Anuradhani Kasturiratne, Shamila Thivanshi De Silva, Anuradha Supun Dassanayaka, Arjuna Priyadarshin De Silva, Norihiro Kato, Arunasalam Pathmeswaran, Ananda Rajitha Wickramasinghe, Hithanadura Janaka de Silva

## Abstract

**Introduction:** Metabolic (dysfunction)-associated fatty liver disease (MAFLD) is a recently suggested alternative to Non-alcoholic fatty liver disease (NAFLD). We compared baseline metabolic traits and outcomes of NAFLD and MAFLD.

**Methods:** In an ongoing, community-based, cohort study, participants were first screened in 2007 by structured-interview, anthropometry, liver ultrasonography, and biochemical/serological tests and reassessed after 7-years. Baseline characteristics and outcomes after 7-years were compared in NAFLD and MAFLD, in those excluded by the NAFLD definition but captured by the MAFLD definition and those excluded by the MAFLD definition but captured by the NAFLD definition, versus controls.

**Results:** Of 2985 recruited in 2007, 940 (31.5%) had NAFLD, 990 (33.1%) had MAFLD and 362 (12.1%) were controls. Compared to NAFLD, MAFLD captured an additional 2.9% of individuals from the cohort and lost 1.3%. At baseline, anthropometric and metabolic traits were similar in NAFLD and MAFLD. At follow-up after 7 years, the odds of having new-onset metabolic traits and fatal/non-fatal CVEs were similar in the two groups, but were significantly higher in both the groups compared to controls. However, those excluded by the NAFLD definition but captured by the MAFLD definition, showed higher baseline metabolic derangements compared to those excluded by the MAFLD definition but captured by the NAFLD definition and had higher odds for having new onset metabolic traits and CVEs compared to controls.

**Conclusion:** Even though it was able to increase the index population by only a small proportion, redefining NAFLD as MAFLD seemed to improve clinical utility.

**Lay Summary:**

- The term “metabolic (dysfunction)-associated fatty liver disease (MAFLD)” has recently been suggested as an alternative to “non-alcoholic fatty liver disease (NAFLD)”.
- In our study, the diagnostic criteria for MAFLD detected a slightly higher number of individuals than NAFLD from the community.
- The proportions of anthropometric and metabolic abnormalities in NAFLD and MAFLD were similar. The adverse outcomes of NAFLD and MAFLD were also similar, but were worse than in a population without NAFLD or MAFLD.
- Those who were excluded by the NAFLD definition and captured by the MAFLD definition seem to be at higher risk of adverse outcomes than those excluded by the MAFLD definition but captured by the NAFLD definition.
- Although it was able to increase the index population by only a small proportion, redefining NAFLD as MAFLD seems useful.

## Introduction

The term non-alcoholic fatty liver disease (NAFLD) was introduced nearly 35 years ago (1).NAFLD is defined by the presence of hepatic steatosis (affecting at least 5% of hepatocytes), detected on imaging or histology, in individuals who consume safe limits or no alcohol, and who do not have a secondary cause of hepatic steatosis (such as viral hepatitis, steatogenic medications or lipodystrophy) (2). NAFLD has evolved into an umbrella term that comprises a spectrum of fatty liver disease. The disease spectrum includes simple, non-alcoholic fatty liver (NAFL) – the potentially non-progressive sub-type, as well as advanced non-alcoholic steatohepatitis (NASH) – the potentially progressive subtype that can lead to severe fibrosis and cirrhosis resulting in liver-related morbidity and mortality (3-6). The NAFLD terminology seems, so far, to have stood the test of time.

The adverse outcomes of NAFLD are associated with the degree of hepatic fibrosis and being metabolically active (7-12). Metabolic dysfunction is also commonly seen in patients with fatty liver regardless of alcohol use (13), and the acronym NAFLD does not accurately reflect this association. Therefore, the more inclusive term metabolic (dysfunction) associated fatty liver disease (MAFLD) has been suggested as being more appropriate (14, 15). There is an ongoing debate as to whether redefining NAFLD as MAFLD would be meaningful (16, 17). A change in nomenclature needs to carefully consider a number of relevant issues, of which long-term outcome is possibly the most important.

The Ragama Health Study (RHS) is a relatively large, community-based study on non-communicable diseases (18). It is a collaborative study between the National Centre for Global Health and Medicine, Tokyo, Japan and the Faculty of Medicine, University of Kelaniya, Ragama, Sri Lanka. In this ongoing, prospective cohort study, we compared anthropometric/metabolic outcomes and fatal and non-fatal cardiovascular events (CVE) between NAFLD and MAFLD compared to controls, after 7-years of follow-up.

## Methods

### Study setting and population

The study population was initially selected from the Ragama, Medical Officer of Health (MOH) area, Sri Lanka, situated 18 km north of the commercial capital, Colombo. The geographical extent of Ragama MOH area is 25 km^2^. The population is urban and multi-ethnic. There were 75,591 people with 15,137 housing units in 2007. The Ragama MOH area is divided into 22 Grama Niladhari (GN) divisions, the smallest administrative units in the country. The householders list of each GN division was used for stratified random sampling. Those aged 35–64 years were identified and stratified into three age groups: 35–44, 45–54 and 55–64 years. A list of all individuals in each stratum was prepared and enumerated. A random sample was obtained from each stratum using a random numbers generator (PEPI statistical program). A sample of 40 individuals from 35–44-year olds and 80 each of 45–54 and 55–64-year olds were identified for selection from each GN division (18). All selected individuals were visited at their homes and invited to participate. The purpose of the study, procedures to be carried out and potential hazards and benefits of participation were explained, prior to obtaining informed, written consent.

### Screening at baseline in 2007 and follow up in 2014

Participants were screened by structured interview. The target population screened initially in 2007 was invited back after 7-years of follow-up for re-evaluation in 2014. Measurement of anthropometric indices, liver ultrasonography, and biochemical and serological tests was done on both occasions. Participants with abnormal anthropometric and metabolic derangements were referred for medical care whenever these were detected at baseline and follow-up. Details of the screening process of the inception cohort in 2007 and re-evaluation in 2014 are described elsewhere (18, 19).

NAFLD was diagnosed on established ultrasound criteria (20) (two out of the following three criteria: increased echogenicity of the liver compared to kidney and spleen, obliteration of the vascular architecture of the liver and deep attenuation of the ultrasonic signal), safe alcohol consumption (Asian standards: <14 units/week for men, <7 units/week for females) (21) and absence of Hepatitis B and C markers (2). MAFLD was diagnosed on the proposed, consensus recommendation (14, 15): ultrasound criteria for fatty liver with (i) a body mass index (BMI)>=23 kg/m2 or diabetes (fasting blood sugar (FBS)>125 mg/dL or HbA1c >6.4% or on treatment), or (ii) if BMI <23 kg/m2 and not diabetic, with at least two other features of metabolic dysregulation, namely, waist circumference (WC) >=90cm for men, WC >=80cm for women; blood pressure (BP)>=130/85 mmHg or on treatment; triglycerides (TG)>150mg/dL or on treatment; high density lipoproteins (HDL)<40mg/dL for men and <50 mg/dL for women or on treatment; and FBS between 100-125 mg/dL or HbA1c 5.7-6.4%. Controls were defined as those who had none of the three ultrasound criteria for fatty liver, no unsafe alcohol use, no diabetes, BMI <23 kg/m2, and less than two of the other features of metabolic dysregulation.

### Follow-up for cardiovascular disease (CVD) outcomes in 2014

At re-evaluation in 2014, participants now aged 42-71-years, were also assessed for fatal and non-fatal cardiovascular events (CVE). CVEs were confirmed by examination of the relevant medical records/death certificates. Non-responders for re-evaluation in 2014 were contacted either by telephone or by post. After obtaining verbal consent, the participant or a collateral historian, in case the participant was deceased, was interviewed by telephone. Details regarding whether the study participant was alive or dead, and if alive, whether the participant had suffered any non-fatal CVEs during the follow-up period were obtained. These CVEs were confirmed during home visits by medically trained research assistants, based on examination of death certificates and hospital records.

### CVE endpoints

Non-fatal CVEs were identified either by documentation in medical records or, in the absence of medical records, by the presence of an episode with clinically significant chest pain or clinically significant neurological deficit, or intervention for either (22). Clinically significant chest pain was defined as chest pain requiring hospital admission for more than 24 hours and requiring special treatment (streptokinase, heparin, cardiology referral with or without echocardiography and/or referral for coronary intervention). Clinically significant neurological deficit was defined as a sudden onset neurological deficit lasting more than 24 hours, requiring a CT scan of the brain and/or management in a specialized Stroke Unit.

Endpoints assessed were fatal-CVE [death from myocardial infarction or stroke] and non-fatal CVE [non-fatal myocardial event (NFME), non-fatal stroke (NFS), coronary artery bypass surgery (CABS), and percutaneous transluminal coronary angiography (PTCA)].

### Lean-NAFLD and lean-MAFLD

We also investigated a sub-population of lean individuals in the RHS cohort. Lean was defined as a BMI<23 kg/m^2^. Based on this, we defined lean-NAFLD as those with NAFLD and BMI<23 kg/m^2^ and lean-MAFLD as those with MAFLD and BMI<23 kg/m^2^.

### Data analysis

Data were entered in Epi Info 7 (Centers for Disease Control and Prevention, Atlanta, GA, USA). Logical and random data entry checks were done. Continuous and categorical data were described using median with interquartile ranges and frequencies with percentages, respectively. Group comparisons were done using Wilcoxon Rank Sum test and the two sample test for proportions. P<0.05 was considered significant.

We initially compared the percentages classified as NAFLD and MAFLD among the original cohort from 2007.We then compared anthropometric and metabolic outcomes, and CVEs between NAFLD and MAFLD and compared them with controls after 7-years of follow-up. Similarly, we compared these parameters between those excluded by the NAFLD definition but captured by the MAFLD definition and those excluded by the MAFLD definition but captured by the NAFLD definition, and compared them with controls after 7-years follow-up. We also compared anthropometric and metabolic outcomes and CVEs between lean-NAFLD and lean-MAFLD and compared them to controls after 7-years of follow-up.

We considered new onset general obesity, central obesity, diabetes, hypertension, hypertriglyceridemia, low HDL and CVEs as outcomes after 7 years of follow up. Unadjusted odds ratios for the occurrence of these outcomes were obtained from simple binomial logistic regression models. These outcomes were further evaluated after adjusting for age, sex and the respective variable of interest at baseline using multiple logistic regression models and were presented as adjusted odds ratios. All the odds ratios were presented with 95% confidence intervals. The analysis was performed using R programming language version 3.6.3 (R Core Team (2020). R: A language and environment for statistical computing. R Foundation for Statistical Computing, Vienna, Austria).

### Ethical considerations

Ethical approval for the study was obtained from the Ethics Review Committee of the Faculty of Medicine, University of Kelaniya. Informed written consent was obtained from all participants in the RHS at baseline and at follow up. Verbal and implied consent was obtained from respondents who did not attend follow up in 2014 but were traced later.

## Results

Of the 2985 recruited in the inception cohort in 2007, 940 (31.5%) had NAFLD [617 (65.6%) women; median-age 53.0 (IQR : 48-59) years], 990 (33.2%) fulfilled criteria for MAFLD [610 (61.6%) women; median-age 53.0 (IQR : 48-59) years] and 362 (12.1%) were controls [193 (53.3%%) women; median-age 50.0 (IQR : 45-58) years] (Table 1).

**Table 1:** Characteristics NAFLD and MAFLD from the original cohort (2007)

| **Prevalence in 2007** | **NAFLD**<br>N = 940 | **MAFLD**<br>N = 990 | **P value** |
| --- | --- | --- | --- |
| Median age (IQR) | 53.0 (48.0 – 59.0) | 53.0 (48.0 – 59.0) | 0.82 |
| Female sex | 617/940 (65.6%) | 610/990 (61.6%) | 0.07 |
| General obesity | 644/936 (68.8%) | 699/989 (70.7%) | 0.37 |
| Central obesity | 792/940 (84.3%) | 850/990 (85.9%) | 0.32 |
| DM | 275/939 (29.3%) | 293/989 (29.6%) | 0.87 |
| HTN | 525/940 (55.9%) | 567/990 (57.3%) | 0.53 |
| Raised TG | 449/940 (47.8%) | 483/990 (48.8%) | 0.65 |
| Low HDL | 310/940 (33.0%) | 318/990 (32.1%) | 0.69 |

When compared to NAFLD, MAFLD captured an additional 88 (2.9%) individuals of the population (fatty liver with metabolic abnormality and alcohol use), and lost 38 (1.3%) (lean-NAFLD without metabolic dysregulation) (Figure 1).

**Figure 1:**
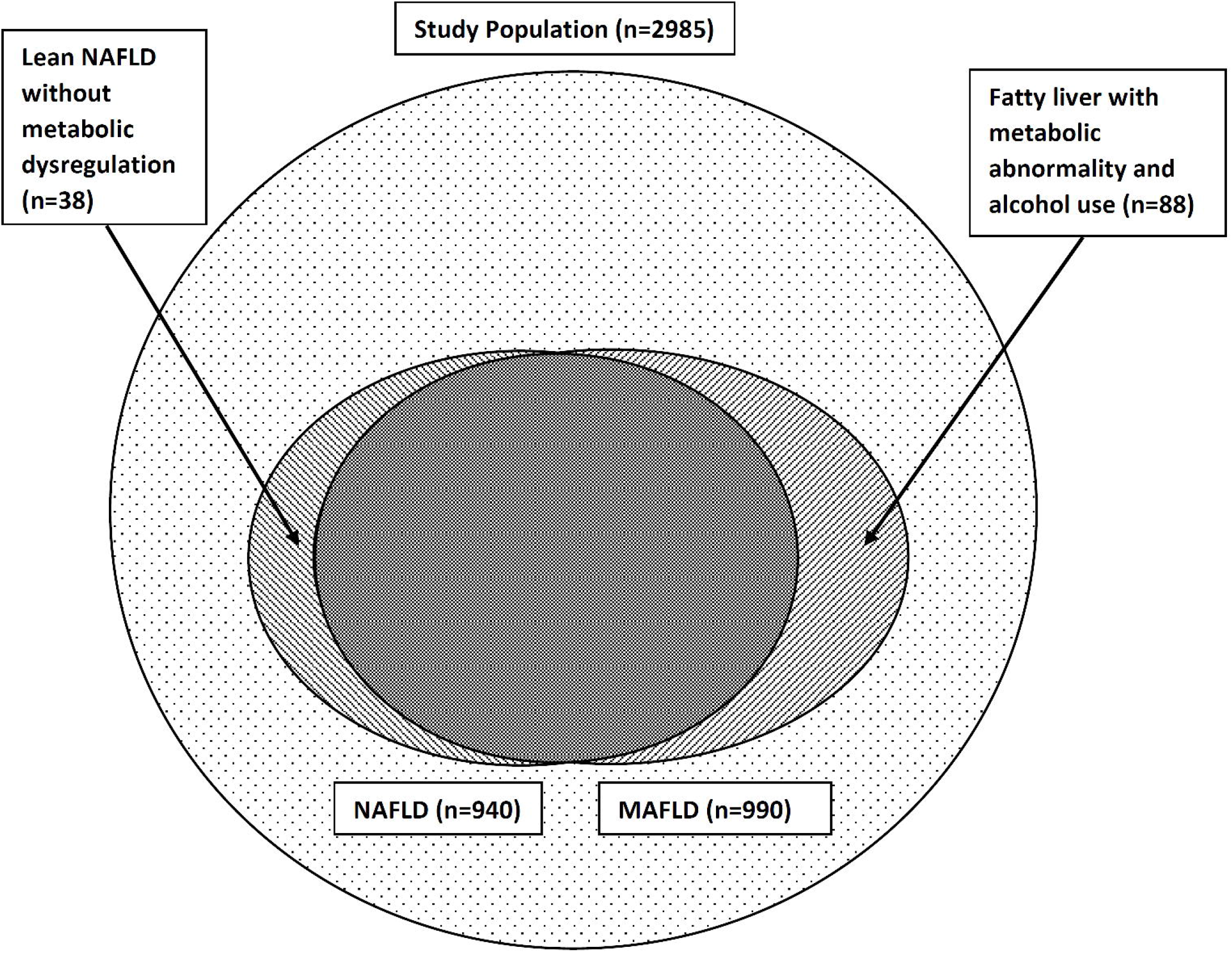
MAFLD, NAFLD and those excluded by the two definitions in the original study population.

2148 (71.9%) (1237[57.6%] women, mean age-59.2 years [SD-7.7]) attended follow-up in 2014. Except for fewer males attending follow up, rest of the characteristics were similar among the inception (2007) and follow up (2014) cohorts (19). The follow-up cohort included 708 who were classified as having NAFLD [472 (66.7%%) women; median-age 54.0 (IQR : 48.0 – 58.2) years], 735 who were classified as having MAFLD [462 (62.9%) women; median-age 53.0 (IQR : 48.0 – 58.5) years] and 255 controls [144 (56.5%) women; median-age 51.0 (IQR : 45.0 – 59.0) years] at baseline in 2007.

At baseline (in 2007), anthropometry and metabolic traits were similar in NAFLD and MAFLD (Table 1). At follow-up after 7 years (in 2014), the odds of having new-onset metabolic traits and fatal/non-fatal CVEs were similar in the two groups, but were significantly higher in both groups compared to controls (Tables 2 and 3).

**Table 2:** New-onset metabolic features and cardiovascular events (with n, %) after 7-years (2014), among control, NAFLD and MAFLD.

| **Outcomes in 2014** | **Controls**<br>N = 255 | **NAFLD**<br>N = 708 | **MAFLD**<br>N = 735 |
| --- | --- | --- | --- |
| Incident general obesity | 9/254 (3.5%) | 53/705 (7.5%) | 57/735 (7.8%) |
| Incident central obesity | 39/254 (15.4%) | 37/708 (5.2%) | 44/735 (6.0%) |
| Incident DM | 31/243 (12.8%) | 203/689 (29.5%) | 216/716 (30.2%) |
| Incident HTN | 36/255 (14.1%) | 109/708 (15.4%) | 111/735 (15.1%) |
| Incident TG | 68/245 (27.8%) | 145/695 (20.9%) | 153/723 (21.2%) |
| Incident low HDL | 68/248 (27.4%) | 246/695 (35.4%) | 250/723 (34.6%) |
| CVD non-fatal and fatal events | 4/255 (1.6%) | 43/708 (6.1%) | 50/735 (6.8%) |

**Table 3:** New-onset metabolic features and cardiovascular events after 7-years (2014) [with OR (95% CI] among NAFLD and MAFLD compared to controls.

| **Outcomes in 2014** | **Exposure variable** |  |  |  |
| --- | --- | --- | --- | --- |
|  | **Unadjusted OR**<br>**for**<br>**NAFLD vs**<br>**controls** | **Adjusted OR**<br>**for**<br>**NAFLD vs**<br>**controls** | **Unadjusted OR**<br>**for**<br>**MAFLD vs**<br>**controls** | **Adjusted OR**<br>**for**<br>**MAFLD vs**<br>**controls** |
| Incident general obesity | 2.2 (1.1 - 4.6) | 10.7 (5.1 – 22.7) | 2.3 (1.1 – 4.7) | 12.1 (5.7 – 25.4) |
| Incident central obesity | 0.3 (0.2 – 0.5) | 4.5 (2.6 – 7.7) | 0.4 (0.2 – 0.6) | 6.8 (4.0 – 11.7) |
| Incident DM | 2.9 (1.9 – 4.3) | 4.5 (3.0 – 6.9) | 3.0 (2.0 – 4.4) | 4.7 (3.1 – 7.2) |
| Incident HTN | 1.1 (0.7 – 1.7) | 2.6 (1.8 – 3.8) | 1.1 (0.7 – 1.6) | 2.6 (1.8 – 3.7) |
| Incident TG | 0.7 (0.5 – 0.9) | 1.5 (1.1 – 2.1) | 0.7 (0.5 – 0.9) | 1.6 (1.2 – 2.3) |
| Incident low HDL | 1.5 (1.1 – 2.0) | 2.5 (1.7 – 3.5) | 1.4 (1.1 – 1.9) | 2.6 (1.8 – 3.7) |
| CVD event - non-fatal +<br>fatal | 4.1 (1.4 – 11.4) | 3.9 (1.4 – 11.2) | 4.5 (1.7 – 12.8) | 4.5 (1.6 – 12.8) |

Those excluded by the NAFLD definition but captured by the MAFLD definition showed higher baseline metabolic derangements (in 2007), except for low HDL, compared to those excluded by the MAFLD definition but captured by the NAFLD definition (Table 4). At follow-up after 7 years (in 2014), those excluded by the NAFLD definition but captured by the MAFLD definition had higher odds for developing incident general obesity, central obesity, DM and CVE compared to controls (Tables 5 and 6). Those excluded by the MAFLD definition but captured by the NAFLD definition only had higher odds for developing incident DM compared to controls.

**Table 4:** Characteristics of those excluded by NAFLD and MAFLD definitions from the original cohort (2007)

| **Prevalence in 2007** | **Excluded by**<br>**NAFLD definition**<br>**but captured by**<br>**MAFLD definition**<br>**N = 88** | **Excluded by**<br>**MAFLD definition**<br>**but captured by**<br>**NAFLD definition**<br>**N = 38** | **P value** |
| --- | --- | --- | --- |
| Median age (IQR) | 51.0 (46.0 – 58.0) | 53.5 (49.0 – 58.0) | 0.477 |
| Female sex | 9/88 (10.2%) | 16/38 (42.1%) | 0.441 |
| General obesity | 58/88 (65.9%) | 3/35 (8.6%) | <0.001 |
| Central obesity | 65/88 (73.9%) | 7/38 (18.4%) | <0.001 |
| DM | 20/88 (22.7%) | 2/38 (5.3%) | 0.0178 |
| HTN | 51/ 88 (57.9%) | 9/38 (23.7%) | <0.001 |
| Raised TG | 44/88 (50.0%) | 10/38 (26.3%) | 0.014 |
| Low HDL | 15/88 (17.0%) | 7/38 (18.4%) | 0.852 |

**Table 5:** New-onset metabolic features and cardiovascular events (with n, %) after 7-years (2014) among controls and those excluded by NAFLD and MAFLD definitions.

| **Outcomes in 2014** | **Controls**<br>**N = 255** | **Excluded by**<br>**NAFLD definition**<br>**but captured by**<br>**MAFLD definition**<br>**N = 57** | **Excluded by**<br>**MAFLD definition**<br>**but captured by**<br>**NAFLD definition**<br>**N = 30** |
| --- | --- | --- | --- |
| Incident general obesity | 9/254 (3.5%) | 5/57 (8.8%) | 1/27 (3.7%) |
| Incident central obesity | 39/254 (15.4%) | 11/57 (19.3%) | 4/30 (13.3%) |
| Incident DM | 31/243 (12.8%) | 21/56 (37.5%) | 8/29 (27.6%) |
| Incident HTN | 36/255 (14.1%) | 8/57 (14.0%) | 6/30 (20.0%) |
| Incident TG | 68/245 (27.8%) | 11/57 (19.3%) | 3/29 (19.3%) |
| Incident low HDL | 68/248 (27.4%) | 10/57 (17.5%) | 6/29 (20.7%) |
| CVD non-fatal and fatal events | 4/255 (1.6%) | 8/57 (14.0%) | 1/29 (3.3%) |

**Table 6:** New-onset metabolic features and cardiovascular events after 7-years (2014) [with OR (95% CI] among those excluded by NAFLD and MAFLD definitions compared to controls.

| **Outcomes in 2014** | **Exposure variable** |  |  |  |
| --- | --- | --- | --- | --- |
|  | **Excluded by NAFLD definition but captured by MAFLD definition vs controls - unadjusted OR** | **Excluded by NAFLD definition but captured by MAFLD definition vs controls - adjusted OR** | **Excluded by MAFLD definition but captured by NAFLD definition vs controls - unadjusted OR** | **Excluded by MAFLD definition but captured by NAFLD definition vs controls - adjusted OR** |
| Incident general obesity | 2.6 (0.8 – 8.1) | 5.3 (1.5 – 18.8) | 1.1 (0.1 – 8.6) | 1.2 (0.1 – 9.6) |
| Incident central obesity | 1.3 (0.6 – 2.8) | 33.2 (8.6 – 127.9) | 0.8 (0.3 – 2.6) | 1.4 (0.4 – 4.7) |
| Incident DM | 4.1 (2.1 – 7.9) | 6.5 (2.9 – 14.4) | 2.6 (1.1 – 6.4) | 2.8 (1.1 – 7.0) |
| Incident HTN | 1.0 (0.4 – 2.3) | 2.1 (0.8 – 5.5) | 1.5 (0.6 – 4.0) | 1.6 (0.6 – 4.3) |
| Incident TG | 0.6 (0.3 – 1.3) | 1.3 (0.6 – 2.9) | 0.3 (0.1 – 1.0) | 4.2 (0.1 – 1.4) |
| Incident low HDL | 0.6 (0.3 – 1.2) | 2.0 (0.8 – 5.1) | 0.7 (0.3 – 1.8) | 9.4 (0.3 – 2.9) |
| CVD event - non-fatal + fatal | 10.2 (3.0 – 35.4) | 8.5 (2.2 – 32.8) | 2.2 (0.2 – 20.0) | 2.0 (0.2 – 19.2) |

There were 121 participants with lean-NAFLD, 98 with lean-MAFLD and 362 controls at baseline in 2007. Of these individuals, 85 lean-NAFLD and 68 lean-MAFLD presented for follow-up in 2014. Anthropometric and metabolic outcomes among lean-NAFLD and lean-MAFLD are given in Supplementary Table 1. The odds of having fatal/non-fatal CVEs were similar in Lean-NAFLD and Lean-MAFLD, but were significantly higher compared to controls. However, the odds of having adverse anthropometric and metabolic outcomes in the two groups compared to controls were not consistent (Supplementary Table 2).

## Discussion

The prevalence of MAFLD and NAFLD in the RHS inception cohort were 33.1% and 31.5%, respectively. When compared to NAFLD, MAFLD captured an additional 2.9% of individuals from the cohort (fatty liver with metabolic abnormality and/or alcohol use), and lost 1.3% (NAFLD with no obesity or metabolic abnormality). At baseline, anthropometric and metabolic traits were similar in NAFLD and MAFLD. At follow-up in 7 years, the odds of having new-onset metabolic traits and fatal/non-fatal CVEs were similar in the two groups, but were significantly higher in both the groups compared to controls. However, those who were excluded by the NAFLD definition but captured by the MAFLD definition showed higher baseline metabolic derangements, except for low HDL, compared to those excluded by the MAFLD definition but captured by the NAFLD definition. At follow-up after 7 years they also had higher odds for developing incident general obesity, central obesity, DM and CVE compared to controls, while those excluded by the MAFLD definition but captured by the NAFLD definition only had higher odds for developing incident DM compared to controls. Therefore, those excluded by the NAFLD definition and captured by the MAFLD definition seem to be a higher risk population than those excluded by the MAFLD definition but captured by the NAFLD definition. This provides support for the argument to change the nomenclature and the definition of fatty liver from NAFLD to MAFLD (17).

We attempted a similar analysis of a subpopulation of lean individuals in the RHS cohort. As the numbers were small, the results of this subgroup analysis (Supplementary Tables 1 and 2) should be interpreted with caution.

The basis of the proposal to change NAFLD to MAFLD is that NAFLD is a diagnosis of exclusion, with prominence given to excluding unsafe alcohol in its aetiology with no provision made for the recognition of the commonly associated metabolic dysfunctions. In contrast, MAFLD is a diagnosis of inclusion and incorporates all potentially relevant metabolic dysfunctions associated with fatty liver, irrespective of alcohol use or pre-existing liver disease (14, 15). However, more recently, there has been debate and a call for caution against a “premature”, widespread adoption of the new definition without careful assessment of the implications of such a change (16). We also believe that any change in nomenclature should result in the inclusion of a significant number of additional individuals with fatty liver from the population, and demonstrate a difference in clinical outcome. In our study, the MAFLD definition increased the index population only by a small proportion. However, the MAFLD definition identified and included a higher risk population in the community than the NAFLD definition.

There have been a few recent studies that have adopted the MAFLD definition in defining fatty liver. In a study conducted in Xinxiang, China, the reported age-adjusted prevalence of MAFLD was 29.85% (23). In another study utilizing data from the third National Health and Nutrition Examination Surveys (NHANES) of the United States, MAFLD was diagnosed in 29.7% participants compared to 33.2% with NAFLD in the overall population (24). These prevalence rates are compatible with those observed in the present study, although unlike in that study, we did not find an age difference between NAFLD and MAFLD. In the study by Lin et al., MAFLD definition was presumed to be more practical for identifying patients with fatty liver disease with a high risk of disease progression, based on the presence of hepatic fibrosis (24). However, this was a cross sectional study, with no follow-up outcome data.

To our knowledge this is the first longitudinal study to examine the clinical utility of the proposed MAFLD definition in a real-world setting. The strengths of the study include the large number of participants in whom the data was collected prospectively at baseline in 2007 and at follow up in 2014; more than 70% of study participants attended follow-up after 7 years; the diagnosis of fatty liver was based on well-established ultrasound criteria rather than surrogate markers such as elevated liver enzymes which are known to be less sensitive. However, the lack of information regarding hepatic fibrosis in this cohort and, consequently, the inability to assess liver-related outcomes at follow up is a drawback of our study. There were several other limitations as well: although liver ultrasound was carried out by three trained doctors, inter-observer variability between ultra-sonographers was not formally assessed; information on alcohol consumption was obtained only by direct questioning of participants which may have led to under-reporting of alcohol use with consequent overestimation of the prevalence of NAFLD.

## Conclusion

In our study, individuals with NAFLD and MAFLD had similar metabolic traits at baseline. NAFLD and MAFLD also had similar outcomes after 7 years. However, those who were excluded by the NAFLD definition and captured by the MAFLD definition seem to be at higher risk of adverse metabolic outcomes and CVEs than those excluded by the MAFLD definition but captured by the NAFLD definition. Thus, although it was able to increase the index population by only a small proportion, redefining NAFLD as MAFLD seems to improve clinical utility.

However, more evidence is required from larger, longer-term outcome studies before strong recommendations can be made to replace NAFLD with MAFLD in clinical practice.

## Data Availability

Data related the the paper will be made available on request to the Ethics Review Commitee of the Faculty of Medicine, University of Kelaniya.

## List of abbreviations

NAFLD: Non-Alcoholic Fatty Liver Disease
NAFL: Non-Alcoholic Fatty Liver
NASH: Non-Alcoholic Steatohepatitis
MAFLD: Metabolic (dysfunction) Associated Fatty Liver Disease
RHS: Ragama Health Study
CVE: cardiovascular events
BMI: Body Mass Index
FBS: fasting blood sugar
WC: waist circumference
BP: blood pressure
TG: triglycerides
HDL: high density lipoproteins
CVD: Cardiovascular Disease
IQR: Interquartile range

## Acknowledgments

We thank those who have continuously supported the Ragama Health Study. We would like to acknowledge the following for their contribution in the data gathering: Withanage KKSA, Goonatillake MDDC, Abeysinghe AACP, De Mel VRT of Faculty of Medicine, University of Kelaniya. We also thank the participants of the Ragama Health Study and the many physicians for their assistance in collecting clinical information.

## Notes

**Conflict of interests** – none

**Financial support:** Grants from the Ministry of Higher Education of Sri Lanka and National Center for Global Health and Medicine, Tokyo, Japan

### Competing Interest Statement

The authors have declared no competing interest.

### Funding Statement

Financial support:
Grants from the Ministry of Higher Education of Sri Lanka and National Center for Global Health and Medicine, Tokyo, Japan

### Author Declarations

Ethics Review Committee of the Faculty of Medicine, University of Kelaniya, Sri Lanka

